# TikTok is an effective platform for bicycle safety injury prevention education

**DOI:** 10.1101/2023.08.02.23293540

**Authors:** Michelina M. Witte, Joey M. McEvoy, Gillian A. Hotz

## Abstract

Social media is an efficient way to spread information, given its widespread daily use by the general population. While it has been shown that public health information can be rapidly disseminated, whether the newer social media platform, TikTok, is effective for this purpose has yet to be explored. The goal of this study is to determine the effectiveness of TikTok to educate about bicycle safety. The TikTok account “iBikeSafe” was created to disseminate injury prevention information related to bicycle safety. Data collected was both qualitative (video classification, comment categorization) and quantitative (likes, views, comments). Performance data was compared between TikTok and another video-sharing social media platform (YouTube). Posts that were didactic (or educational) in nature yielded more views, likes, and comments than posts that were fun or casual in nature. Almost half (46%) of comments made by users on iBikeSafe’s TikTok videos were made to constructively further the discussion and to educate others with accurate injury prevention information. Five of the six videos that were adapted from the iBikeSafe YouTube channel for use on TikTok garnered exponentially more views on TikTok (from 118% to 2057%) than on YouTube, despite having less followers/subscribers (759 TikTok vs. 977 YouTube) and being in existence for less time (11 months vs. 8 years). TikTok is an effective platform for injury prevention education. Didactic posts best provide users with the ability to engage with content and to discuss takeaways, while fun posts keep users active within the platform. Both educational and fun types of videos play an integral role in the effective use of this platform for bicycle safety education. Consideration should be taken into account for the appropriate ratio of didactic vs. fun posts for information dissemination in any public health educational campaign making use of this platform.

## 1. Introduction

Social media has become an established medium for information distribution across all sectors of people’s everyday lives [1]. According to a recent Pew Research report, 48% of Americans under the age of 30 say they go online almost constantly with 46% of them saying they go online multiple times per day [2]. As social media continues to become increasingly entangled in people’s lives, it is important for public health organizations to make appropriate use of relevant social networking platforms to disseminate useful health and safety information and to combat misinformation [3], especially in younger generations whose use of social media has become an integral and ingrained part of their daily routine [4].

Social networking sites have seen a surge in popularity over the past decade. In 2012, there were 1.5 billion collective social media users; that number has since increased by more than 300% to 4.8 billion users as of April 2023 [5]. This means that the extensive reach of social networking sites includes more than half of the current global population of 8 billion people [6]. Given the profound impact and expansive reach of social media, its practical importance to being able to improve public health and to solving health problems has been proposed, along with its advantages and disadvantages [3, 7–9]. Social media provides a new frontier for medicine and healthcare [9], and from the drastic changes brought upon by the COVID-19 pandemic, it has served as an efficient means to widely share information across all ages and ethnic demographics.

TikTok is among the newest social networking sites. After merging with the musical.ly mobile application and launching internationally in 2017, TikTok has gained over 800 million users worldwide with 41% of its users being between the ages of 16 and 24, collectively considered “Generation Z” or “Gen Z” [10]. Reports and studies from a variety of research areas have collectively concluded that the unique style of TikTok – with short, condensed video clips addressing a single objective – is particularly useful in educating this generation [4, 11–13] and has recently been shown to be effective in higher education settings [14, 15]. Given that unintentional injuries are amongst the leading causes of death and disability amongst the younger generations, which saw a significant post-COVID-19 pandemic increase [16], this platform has the potential to reach and educate younger generations with important public health messaging that could mitigate their risk of injury.

While the use of other social media platforms, such as Facebook, Instagram, Twitter and YouTube, for disseminating public health information has been previously considered [8, 17–18], the potential role and usefulness of TikTok for disseminating injury prevention information has yet to be explored. The current study proposes the analysis of TikTok to effectively educate the public about injury prevention, specifically as it relates to bicycle safety. To date, no studies have investigated the potential role of this social media platform as a tool for injury prevention.

The primary goal of this study is to identify the most effective use of TikTok for injury prevention education in the context of bicycle safety. A secondary aim is to determine whether the type of content posted on TikTok, i.e., video posts that are didactic/instructional vs. those that are fun/casual in nature, impacts user engagement and user feedback. The findings from this study will provide guidance on how injury prevention researchers may best make use of this unique, condensed, video clip-sharing social media platform for planning and implementing effective injury prevention education campaigns.

## 2. Methods

### 2.1 Materials

A TikTok account called “iBikeSafe” was created on January 28, 2022. The purpose of establishing the iBikeSafe presence on TikTok was to create and post relevant, innovative injury prevention content to promote the use of bicycles as part of everyday life through advocating for safer bicycle-friendly infrastructure (such as barrier-protected bike lanes), teaching basic bicycle safety skills, and promoting physical activity through bicycling. Two types of video posts were regularly published by iBikeSafe on the TikTok platform: those considered to be fun and those considered to be didactic. Both types of video posts were made regularly and publicly posted to TikTok by iBikeSafe.

### 2.2 Data collection and analysis

All of the data in this study is publicly available, and therefore not subject to ethical review and approval. Performance data and anonymized, collective user data for all of iBikeSafe’s TikTok posts was freely provided by TikTok. The age distribution and the relative percentage of iBikeSafe’s TikTok followers in each age category is broken down as follows: 18-24 years old (32%), 25-34 years old (41%), 35-44 years old (17%), 45-54 years old (6%), 55+ years old (4%). The performance data for iBikeSafe’s video posts was further micro-analyzed using a mixed methods, quantitative and qualitative, approach. The quantifiable parameters of interest from each of the posts include: views, likes, comments, shares, and saves. Also quantified was the relative reach of each of the video posts based on whether each individual post became “highly popular” (meaning it garnered ≥ 4,000 total views) or whether it was “less popular” (meaning it garnered < 4,000 total views). The determination of 4,000 views as the minimum requirement for which video posts were deemed as “highly popular” was made because once all posts were organized in numerical order based on the number of views they received, this threshold emerged as what consistently differentiated a video post with extraordinary popularity vs. one that was not as popular. Moreover, it was observed that the other quantifiable parameters (listed above) exponentially increased once a video reached this threshold.

Qualitative analysis was done on the content posted by TikTok users as comments on iBikeSafe’s video posts. First, a thematic characterization of all user comments that were made on iBikeSafe’s video posts was performed. The top five user comment themes that emerged were: misinformation about bicycle policy/safety, aversion to car-centric city design, furthering discussion and/or educating each other, aggressive anti-bicyclist, and protected bike lane advocacy. Next, each comment sub-category was accounted for by the definitions that follow. *Positive* comments encompass all comments that were in favor of making bicycling part of everyday life, promoting safer bicycle infrastructure, and positive feedback or promotion of the content from the post. *Negative* comments include comments that were against making biking a part of everyday life, promoted car-based infrastructure, and negative feedback and/or demotion of content from the post. *Inquisitive* comments are characterized based on if the comments were asking questions, engaging in discussion through a comment thread, and/or adding more constructive information to what the post was discussing. *Neutral* comments are comments that were: not supporting nor opposing bicycle riding and safety information, not furthering discussion of the topic nor adding more information or asking a question. A selection of actual comments made by TikTok users on iBikeSafe’s posts and their ultimate categorization based on this system can be found in Table 1.

**Table 1.** Examples of comments categorized as positive, negative, inquisitive, and neutral. Table provides examples of actual user comments and locations of where, within the four sub-categories, they were placed.

| Type of Comment | Examples (actual user comments) |
| --- | --- |
| Positive | 1. “We need protective bike lanes” |
|  | 2. "Yes! Bikes and cars do not belong in the same lanes! Give bikes their own infrastructure"<br>3. "Sent to my council members"<br>4. "Wish we had separate and protected bike lanes everywhere"<br>5. "[Protected bike lanes] should be mandatory along every road in America" |
| Negative | 1. "Bikes don't belong in the road"<br>2. "Get a car"<br>3. "Philosophically, why do we want bikers to be alive? I see no need."<br>4. "Bicyclists need to be executed"<br>5. "US cities just aren't bikeable and never will be"<br>6. "Pay taxes to get lanes" |
| Inquisitive | 1. "Which do you prefer, putting pedestrians in danger by speeding down a crowded sidewalk or inconveniencing drivers by making them slow down?"<br>2. "When claiming a lane, you should pull over frequently enough that you never hold more than 5 cars (a typical standard for blocking flow of traffic)."<br>3. "Then maybe cyclists need to pay taxes and then those taxes can pay for infrastructure just like gas tax does for cars?"<br>4. "Roads are largely funded through property taxes and other general local taxes. The idea that drivers alone pay for roads is a myth." |
| Neutral | 1. "These comments make me so sad"<br>2. "Probably Florida"<br>3. "@notjustbikes @pedestriandignity"<br>4. "I got hit the other day, it was terrifying. Wear helmets y'all" |

Lastly, to compare the performance of iBikeSafe’s videos on TikTok vs. its performance on another video sharing social media platform (YouTube), six of the exact same videos were posted both to iBikeSafe’s TikTok account and to its YouTube channel. Table 2 shows the number of views achieved by each video on both of these platforms, as well as subscriber/follower counts and time of iBikeSafe presence on each site.

**Table 2.**
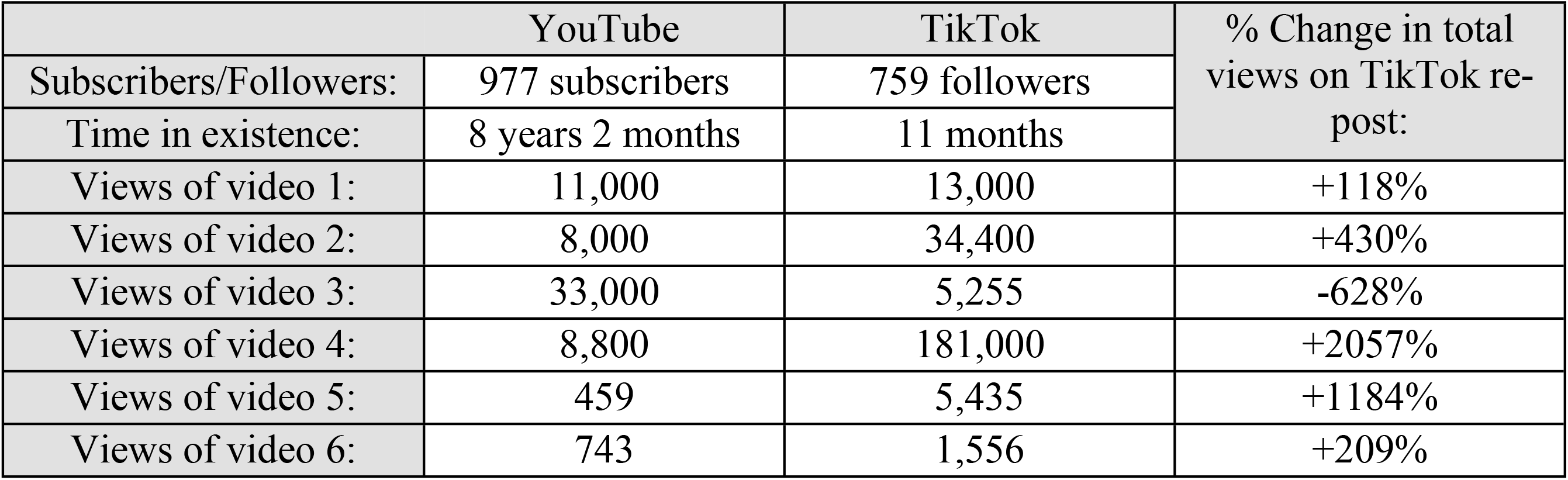
iBikeSafe performance on both YouTube vs. TikTok social media platforms. YouTube videos adapted to TikTok posts led to exponentially greater views (over 100% for five out of six videos and by over 1000% for two out of six).

## 3. Results

On average, iBikeSafe’s fun video posts received 2,226 (± 3,160) views compared to 22,221 (± 53,613) views for didactic video posts (data not shown). Thus, the average views achieved by a didactic video was 898% greater than those achieved by fun videos. Because of the relatively high standard deviations obtained when comparing the totality of video posts, the highly popular (≥ 4,000 views) fun (“X”) and didactic (“Y”) posts were separated from those that were less popular (< 4,000 views) and re-analyzed.

### 3.1 Highly popular posts

Of eleven educational videos and thirty fun videos posted by iBikeSafe, five didactic and five fun videos became highly popular (≥ 4,000 views). The average amount of views, likes, and comments for highly popular fun (“X”) videos compared to highly popular didactic (“Y”) video posts are shown in Fig 1. Highly popular didactic video posts yielded greater views than highly popular fun video posts (47,818 ± 75,395 vs. 8,454 ± 3,774, respectively). Not only did highly popular didactic videos lead to greater views than fun videos, but they also led to 356.4% more likes and 309% more comments than highly popular fun videos. The average number of likes for highly popular fun videos was 910 (± 777) versus 3,243 (± 4217) for highly popular didactic videos. The average number of comments posted by TikTok users on highly popular fun videos was 43 (± 38) versus 176 (±172) posted on highly popular didactic videos. In summary, amongst all highly popular (≥ 4,000 views) video posts, those that were didactic or educational in nature had more views, likes and comments, than those that were more casual and fun.

**Fig. 1.**
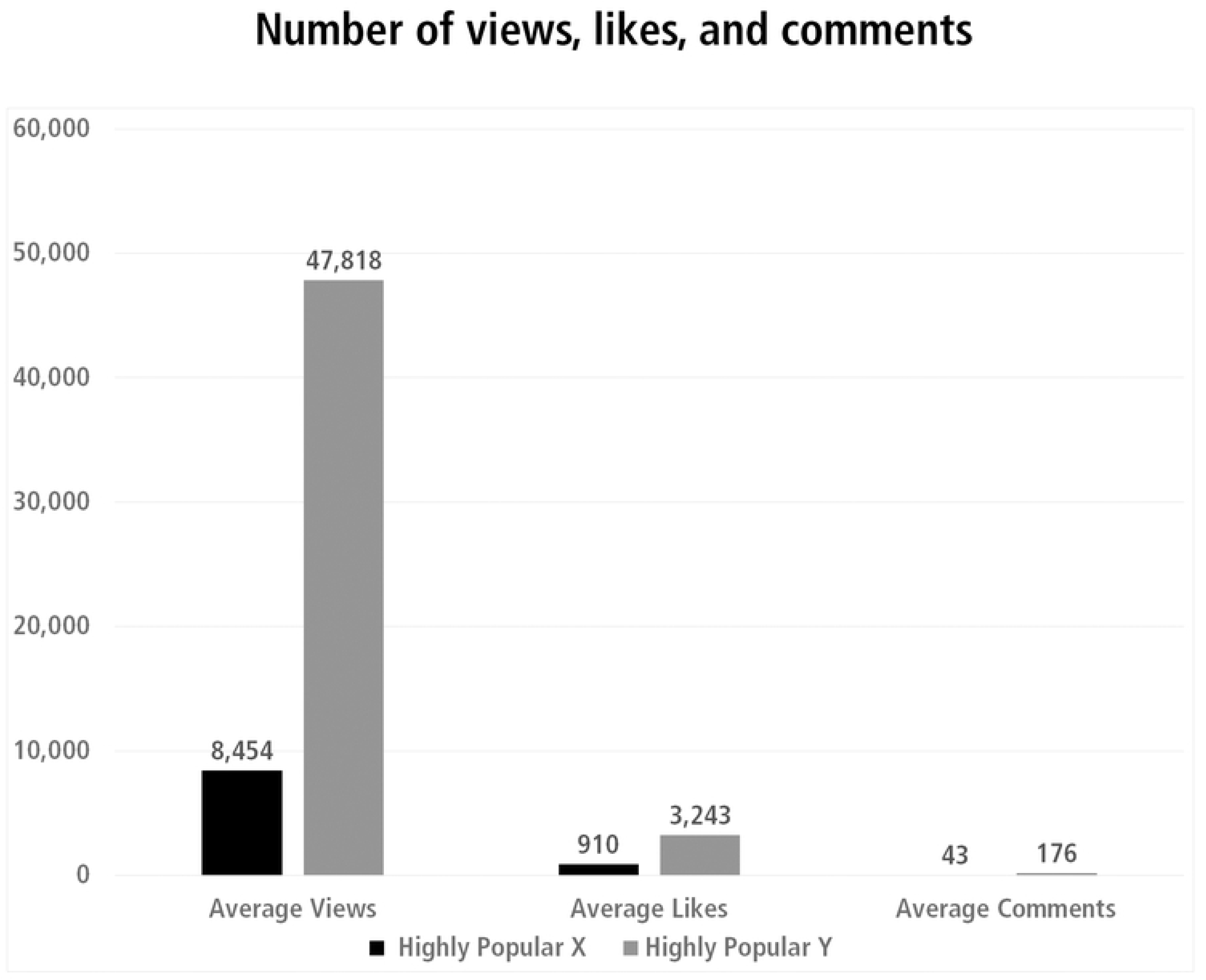
Highly popular fun posts had less views, likes, and comments than highly popular didactic posts. Black bars: fun (“X”) video posts, grey bars: didactic (“Y”) video posts.

### 3.2 Less popular posts

In contrast, the average number of views for less popular fun videos was 1,112 (± 665) versus 890 (± 397) for less popular didactic videos (Fig 2). This indicates that amongst less popular videos (videos that achieved < 4,000 views), those that were fun had 25% more views than those that were didactic. The average number of likes for less popular fun videos was 75 (± 73) versus 54 (± 33) for less popular didactic videos. The average comments made by TikTok users on less popular fun videos was 4 (± 4) while the number of average comments made on less popular didactic videos was 2 (± 1). In all less popular video posts, the fun posts had more views, likes, and comments than the didactic video posts.

**Fig. 2.**
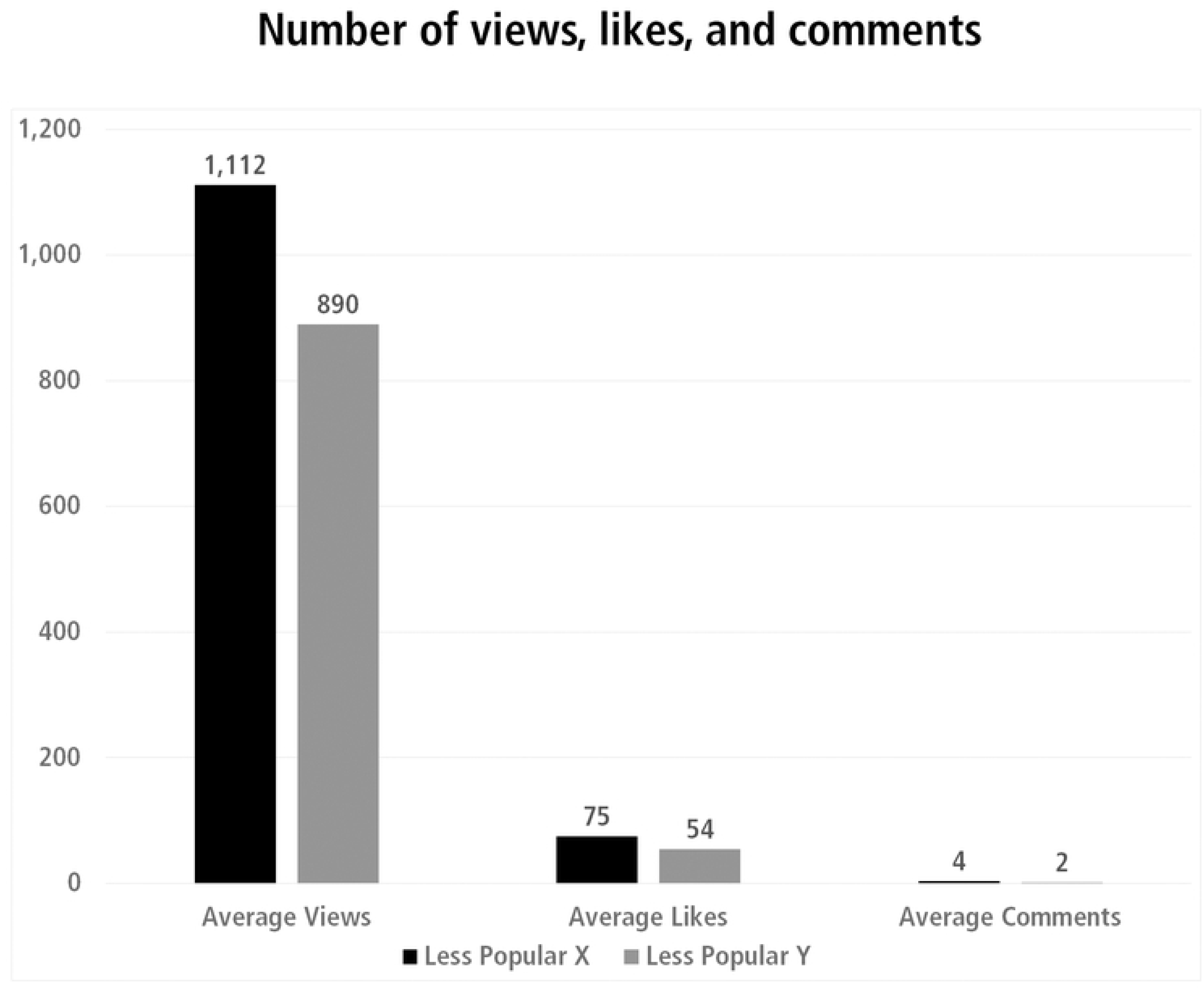
Less popular fun posts had more views, likes, and comments than less popular didactic posts. Black bars: fun (“X”) video posts, grey bars: didactic (“Y”) video posts.

### 3.3 User comments

Amongst the comments made by TikTok users on iBikeSafe’s video posts, five themes emerged (Fig 3). The majority of comments (46%) were made to further the topic of discussion that iBikeSafe’s video post initiated and that led to constructive engagement and education of other TikTok users. Approximately 16% of comments contained misinformation about bicycle safety, but a comparable amount (nearly 15%) of comments were advocating for protected bicycle lanes. Lastly, about 12% of comments expressed anti-bicyclist sentiments, whereas again a comparable amount (11%) of comments were about being averse to car-centric city design.

**Fig. 3.**
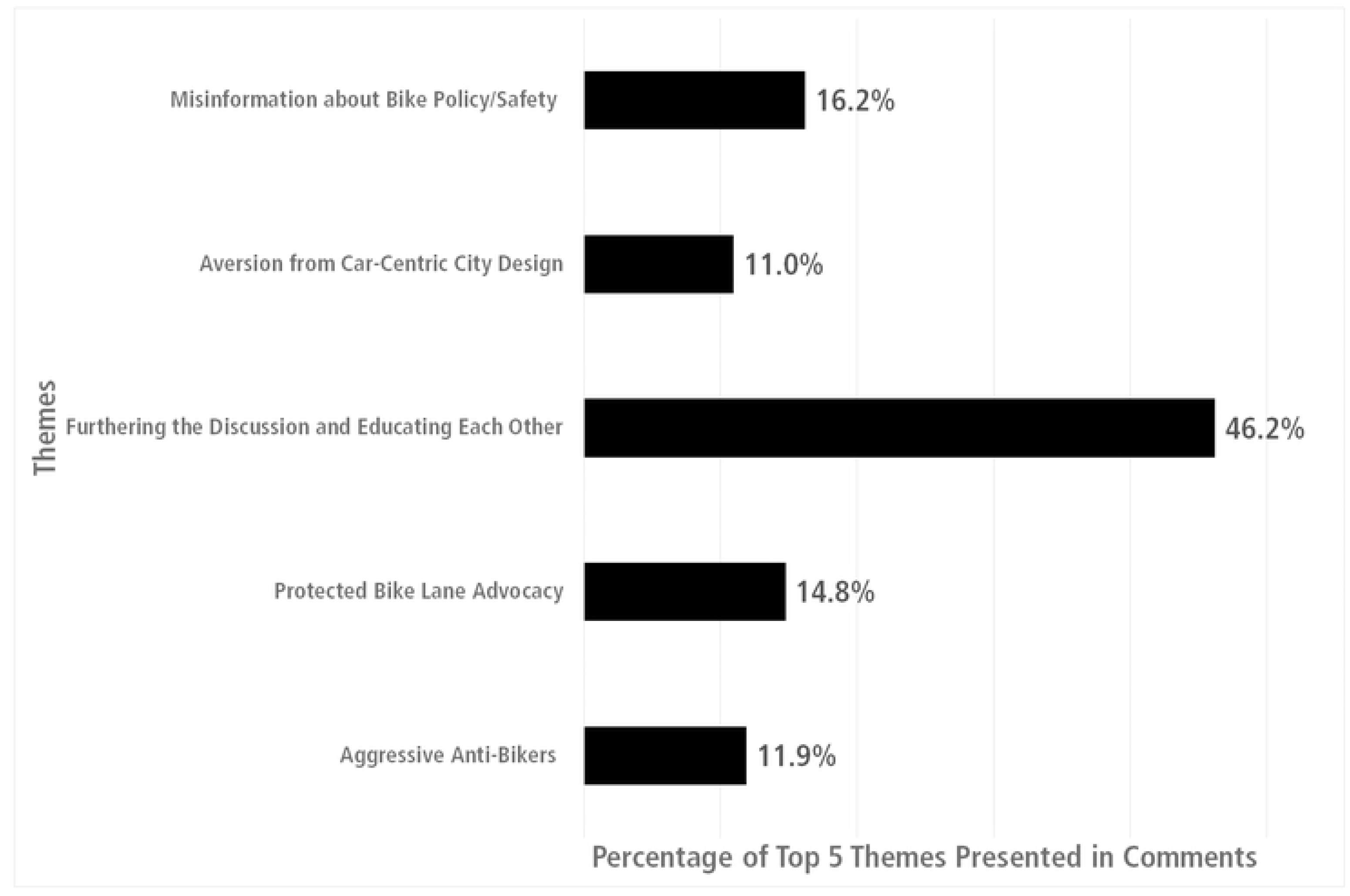
Percentage of each of the five themes that emerged in the comment sections of iBikeSafe’s posts.

TikTok user engagement was further assessed by examining the distribution of positive, negative, inquisitive, and neutral user comments that were made on the top four highly popular didactic (“D”) video posts and the top four highly popular fun (“F”) video posts (Fig 4). While the data show that didactic posts most often led to a greater number of inquisitive user comments, inquisitive user comments were consistently the most frequent type of comment made by TikTok users on iBikeSafe’s posts regardless of post type (didactic vs. fun) and amount of views (most vs. least). This suggests that a combination of both didactic and fun posts can be useful for constructive user engagement on TikTok. A sampling of the actual comments made by TikTok users on iBikeSafe’s posts can be found in Table 1.

**Fig. 4.**
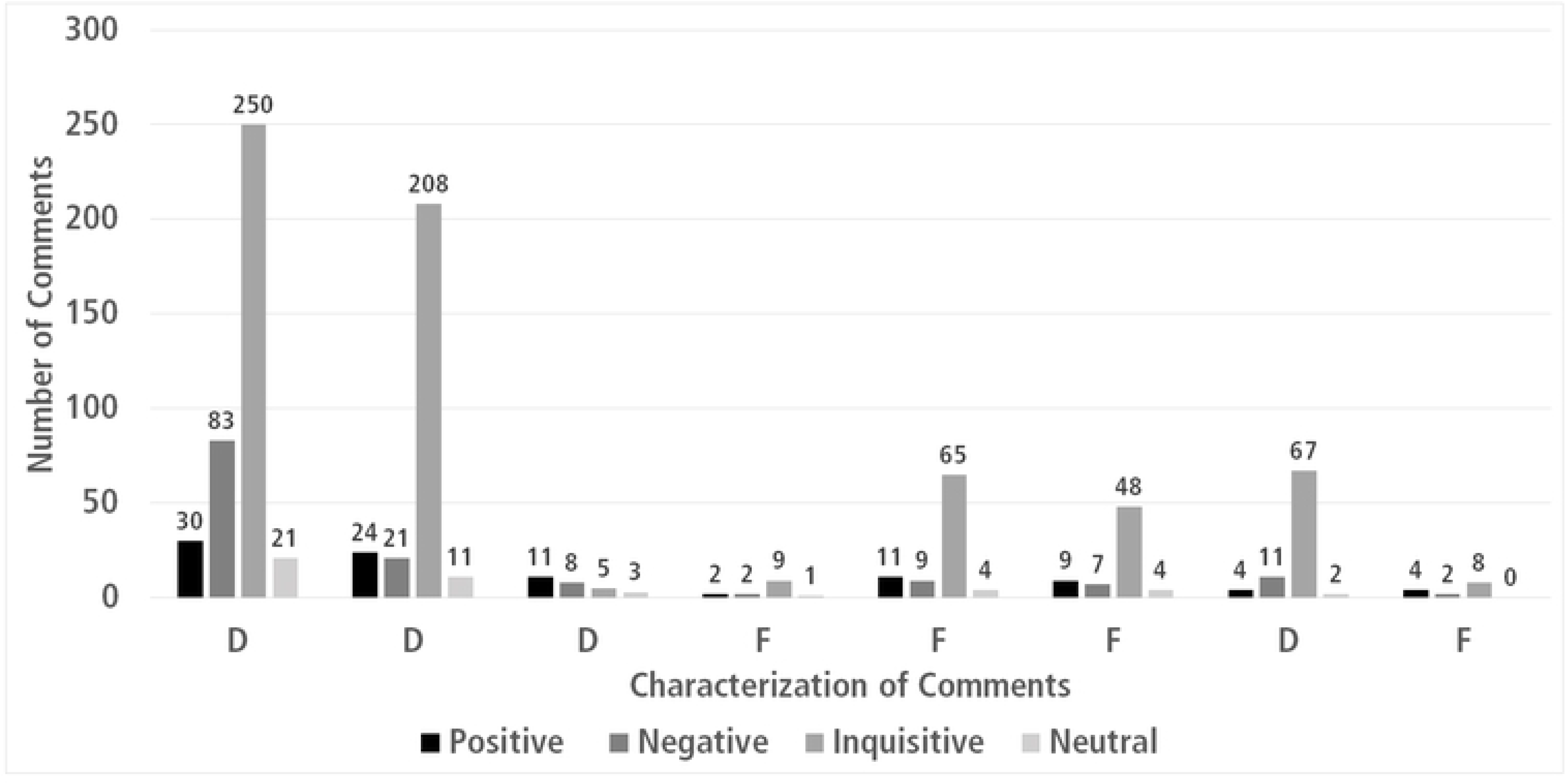
User comments were categorized into four sub-types: positive, negative, inquisitive, and neutral. Black bars: positive user comments, dark grey bars: negative user comments, light grey bars: inquisitive user comments, white bars: neutral user comments.

### 3.4 TikTok vs. YouTube

Lastly, six of iBikeSafe’s YouTube videos (some of which were fun and some of which were didactic) were adapted to be posted to TikTok. Five of the six videos re-posted to TikTok garnered enough views (≥ 4,000) to be categorized as highly popular (Table 2). Five of the six videos also led to exponential increases (118%, 209%, 430%, 1,184%, and 2,057%) in the number of views when posted to TikTok, compared to when posted to YouTube. Only one of the six videos yielded less views on TikTok than it did on YouTube (5,255 vs. 33,000 views, respectively). Collectively, these findings indicate that TikTok tended to outperform YouTube when sharing the exact same video content adapted for viewing on each respective platform.

## 4. Discussion

The primary purpose of this analysis was to determine the most effective use of TikTok as a means to educate the public about bicycle safety. A secondary goal was to determine whether the type of video content (fun vs. didactic) posted to TikTok would play a significant role in user engagement. This information was sought to inform public health organizations and injury prevention programs on best practices with regards to this unique, condensed, video-clip sharing form of social media to maximize impact. The results revealed that both fun and didactic types of video posts are relevant when using this platform. Among posts that became highly popular (≥ 4,000 total views), those that were didactic in nature led to the greatest amount of user engagement, as indicated by views, likes and comments; however, among the less popular posts (< 4,000 total views), those that were fun in nature had greater user engagement than the didactic posts with similar viewership. This suggests that while TikTok users still view the fun and casual posts that they have come to expect because they are typical of this platform, they actually engage more with posts that are didactic and educational in nature. Given that both fun and didactic types of videos were posted to iBikeSafe (in a ratio of 3:1, fun to didactic), it is reasonable to suggest that both video types likely played a role in follower acquisition and in TikTok user engagement. Deliberate consideration of the ratio of fun vs. didactic posts should be taken into account when looking to effectively educate the public using this platform.

Injury prevention programs have traditionally taken the didactic approach (e.g., incorporating the use of educational infographics, statistics, and flow charts) when utilizing social media as part of their information campaigns [3]. Whether this approach would be appropriate for this newer, modern, micro-video clip sharing platform had yet to be determined prior to this study. To this end, iBikeSafe regularly posted educational/didactic videos to its TikTok page, as well casual/fun videos, the latter of which are most common on the TikTok platform [19]. While the fun videos posted by iBikeSafe were not overtly educational, they still informed users about important bicycle safety concepts. This novel approach of inconspicuously educating TikTok users through the use of original, casual and fun video posts that are typical of this platform, proved effective at garnering user attention. However, while fun videos did generate user activity, the results indicate that using a hybrid technique of posting both fun videos, as well as didactic videos (in a ratio of 3:1, respectively), was most effective at achieving maximal views, likes, and comments.

While total views may be an obvious metric for user engagement on social media, the actual content and number of comments made on posts by TikTok users better elucidates the impact made by the posts. User comment analysis allows for the determination of whether users are learning and/or actively discussing the content with other users to and whether they are elaborating on the injury prevention concepts brough forth by the original post. Comments made by users can be demonstrative of an on-going interest in the ideas posted in the videos and they can be indicative of how the content may reflect the perspective of the person who is commenting. The results affirm that the method used by iBikeSafe successfully led to this type of meaningful engagement, given that nearly half of all user comments were categorized under the theme of *Furthering the Discussion and Educating Each Other,* regardless of the type of post (fun or didactic) that provoked the user to comment. One can objectively contend that inquisitive TikTok user comments are desirable (even over positive TikTok user comments) because they are an indication that organic curious discussion is being sparked and genuine questions are being asked about the content. In other words: *learning* is taking place.

The present study is the first to report that TikTok video sub-types play a role in user engagement. Specifically, didactic or educational videos garnered the greatest number of views, likes and comments amongst videos that reached and/or surpassed 4,000 views. Amongst the videos that reached less than 4,000 views, fun videos garnered more views, likes and comments than educational videos with similar viewership. This suggests that both types of videos can be effective at engaging users on this platform. While the most popular educational videos may have accumulated greater reach (i.e., likes, comments and views), the reach of the less widespread videos favored the fun videos. This means that there is a difference in the way in which TikTok users interact with educational versus fun videos and that both types of videos play an integral role in an injury prevention education campaign utilizing this platform for information dissemination.

Despite the many useful findings with regards to the use of TikTok for injury prevention education, this study was not without its limitations. While iBikeSafe did incorporate appropriate hashtags to increase viewership of users searching for relevant posts related to bicycle safety such as “#bikesafety,” “#protectedbikelanes,” and “#safestreets,” there are other features within TikTok that were not used and could still be considered such as the “live,” “story,” and “repost” features, as well as purchasing the promotion of a video post. Also, metrics such as sharing and saving video posts are available from TikTok, but the current study did not analyze those actions. Lastly, the creation of posts that are deliberately designed for solicitation of users to engage with the content was not utilized.

Regarding educating users on this platform, the qualitative data obtained from the user comments made on iBikeSafe’s video posts indicates that public discussion was furthered and genuine inquisitive comments were made both on the most highly popular educational videos and on the less popular fun videos. The fact that organic discussion was sparked by iBikeSafe amongst TikTok users and is displayed publicly in the comments sections of iBikeSafe’s video posts, demonstrates the effectiveness of this platform as an educational tool. More research is needed to determine if the findings in this study can be applied to other injury prevention and/or public health educational campaigns that seek to use this platform.

In summary, this is first study to demonstrate the effective use of TikTok for injury prevention education, specifically regarding bicycle safety. If used effectively, mass social media campaigns can produce positive public health behavior changes and prevent negative health-related behavior changes across large populations [20]. In a 2021 content analysis of the use of the photo- and short-video sharing social media platform, Instagram, for pediatric injury prevention messaging [21], it was emphasized that injury prevention recommendations made on a visual platform should provide matching visual representations of those recommendations. This suggestion was made in order for the posts to be consistent with the core tenet of social cognitive theory – that people learn through observation [22] and to be effectively utilized on a visual platform. Furthermore, in a systematic review on the use of social media for injury prevention [17], it was acknowledged that social media offers the opportunity to influence and steer public perception about injuries and it was suggested that public health entities should use patterns of major news stories to influence online conversations about injury. Future studies should further examine these approaches and recommendations.

## 5. Conclusion

The present study is the first to establish that TikTok can serve as an effective platform for public health messaging, particularly in the realm of injury prevention and bicycle safety. Appropriate and deliberate considerations should be taken to incorporate a mix of fun video posts and didactic video posts to optimize user involvement. The emergence and identification of common themes in user comments can be used to aid policymakers in proposing concepts (e.g., “Vision Zero” policies), transportation agencies in implementing applicable elements (e.g., complete and/or slow streets design proposals) and bicycle safety education programs in informing strategies. Engaging the community in constructive and innovative ways that invite the opportunity for productive discourse and subsequent education, such as that which is provided by TikTok, is the best approach for reducing bicycle-related injuries and fatalities, especially in younger generations.

TikTok accounts that provide a mix of fun/casual videos, alongside more didactic/educational videos, can achieve success in terms of both quantifiable (comments, likes, views) and qualitative (types of comments) user engagement. The key findings of this study highlight the importance of selective incorporation of didactic posts for widespread community engagement on the TikTok platform and for bringing users to, and keeping them constructively active with, an injury prevention page. The incorporation of complementary fun video posts serves to enhance and perpetuate continued user engagement with the page, whereas the selectively sporadic posting of didactic videos leads to constructive and user-led educational discourse (as evidenced in the comment themes that emerged). Thus, the hybrid use of a combination of both educational video posts and fun video posts allows for injury prevention education on bicycle safety topics to be explored in a variety of contexts.

## Data Availability

All relevant data are within the manuscript and its Supporting Information files.

## Acknowledgments

We would like to express our gratitude to Hengyi Ke of the Biostatistics Collaboration and Consulting Core at the University of Miami Miller School of Medicine’s Department of Public Health Sciences for his generous assistance in providing insight regarding the appropriate statistical and data analyses for this study.

## Notes

### Competing Interest Statement

The authors have declared no competing interest.

### Funding Statement

The author(s) received no specific funding for this work.

### Author Declarations

Anonymous analysis of the social media data obtained in this study did not require Institutional Review Board approval.

